# Does Intermittent Nutrition Enterally Normalise hormonal and metabolic responses to feeding in critically ill adults? The DINE-Normal proof-of-concept study

**DOI:** 10.1101/2025.05.30.25328645

**Authors:** Clodagh E Beattie, Borislavova Borislava, Harry A Smith, Michael Ambler, Paul White, Danielle Milne, Aravind V Ramesh, Alexander Ferriman, Thomas Fisher, Charlotte Horsley, Sherena Jackson, Chloe Jubainville, Kate Lobo, Hannah Maxfield, Javier T Gonzalez, James A Betts, Anthony E Pickering, Matt Thomas

## Abstract

**Background and aims:** For intensive care unit (ICU) patients fed via a nasogastric (NG) tube, current guidelines recommend continuous feeding through the day and night. Emerging evidence in healthy individuals shows that NG feeding in an intermittent diurnal pattern promotes phasic hormonal, digestive and metabolic responses vital for effective nutrition, though this has not been studied in the critically ill population. This proof-of-concept study aimed to compare the effect of diurnal intermittent versus continuous enteral feeding on hormonal and metabolic outcomes in ICU patients.

**Methods:** We conducted a single-centre, randomised, open-label trial in the ICU. Adult ICU patients that were anticipated to require NG feeding for >48 hours were randomised to an intermittent diurnal regimen (feeds at 8:00, 13:00 and 18:00), or continuous feeding with equivalent nutritional value, for 48 hours. The primary outcome was peak plasma insulin within 3 hours of delivering the first intermittent feed on the second study day, compared to the same time period in the continuous group. Secondary outcomes included feasibility, tolerability and metabolic profiles.

**Results:** Thirty patients were randomised to intermittent (n=13) or continuous (n=17) feeding. Two patients in the intermittent group were excluded from analysis (1 required jejunal feeding, 1 withdrew consent). Peak plasma insulin concentrations (mean ± SD) were significantly higher in the intermittent group (373 ± 204pmol/L) versus continuous (58 ± 41pmol/L, p<0.001). Peak plasma glucose concentrations (mean ± SD) also differed in the intermittent and continuous groups, respectively (10.6 ± 2.5mmol/L versus 8.4 ± 2.4mmol/L, p=0.04). There were no other statistically significant between-group differences.

**Conclusion:** Intermittent diurnal feeding, compared to continuous feeding, preserves the physiological insulin response in critically ill adults. Both regimens were well tolerated, supporting the need for a larger trial to assess other clinically important patient-centred outcomes.

**Trial Registration:** This trial was registered prospectively at clinicaltrials.gov (study ID NCT06115044).

## Introduction

Approximately 200,000 patients are admitted to critical care units in the UK every year.(1) In line with international guidelines, approximately 50% of these patients are supported with early enteral nutrition via a nasogastric tube, as they will be unable to feed themselves for a prolonged period.(2) Despite known benefits of enteral nutrition in the prevention of catabolism and improved clinical outcomes, the optimal feeding pattern remains uncertain, and nutritional targets are often missed.(3,4) Historically, most critical care units deliver enteral nutrition as a continuous infusion, although this is unphysiological, failing to align with circadian/diurnal rhythms in both human behaviours (i.e. typical meal patterns) and metabolic responses.(5)

Intermittent feeding, where feed is delivered over periods of 20-60 minutes with fasting intervals in between, has been proposed as an alternative to continuous feeding.(6) International guidelines in favour of continuous feeding are largely founded upon widely accepted practice, and an absence of conclusive evidence for the benefits of intermittent feeding in the ICU plus a potential signal from a few small studies that intermittent feeding may increase of gastrointestinal intolerance, aspiration, and dysglycaemia.(7,8) More recent reviews of the evidence have found no significant differences in aspiration; gastric residual volumes and glucose variation, but highlight a trend towards diarrhoea in intermittent feeding and constipation in continuous feeding groups, respectively.(9–11) The studies included in these reviews show considerable heterogeneity in the feeding regimens, such as the inclusion of nighttime feeding in intermittent arms, which may negate any metabolic benefits of a prolonged overnight fast.

There are other, potentially more compelling, benefits to intermittent feeding. Aligning feeding with wake cycles and fasting with sleep (diurnal feeding), may offer metabolic advantages. Timing of feeds can influence circadian regulation, as synchronisation of nutritional intake with central genetic clocks may alter metabolism of nutrients by peripheral tissues.(12,13) Indeed there is evidence that the timing of feeds can act as a zeitgeber in its own right – so aligning the circadian and metabolic rhythms.(14) In the longer term, misalignment of internal clocks increases the risk of immune dysfunction, insulin resistance and cardiovascular disease.(15–18) In addition, intermittent feeding may better support muscle protein synthesis, enhance gut motility, and better maintain glycaemic control and/or insulin sensitivity.(19–22) Importantly, intake of normal meals in healthy subjects promotes pulsatile release of insulin, a process critical for net anabolism; regulation of glucose, lipid and protein metabolism; and skeletal muscle autophagy and glycaemic control: we don’t know if this is also true in critical illness.(23,24) Pragmatically, intermittent feeding may improve mobility by reducing the need to be connected to a feeding line, and reduce the impact of feed interruptions to help achieve nutritional targets more effectively than continuous feeding.(25) In combination, this could have important impacts in improving metabolic, hormonal, circadian and long-term functional outcomes.

The aim of the DINE-Normal study was to evaluate both the feasibility and the metabolic and hormonal effects of diurnal intermittent versus continuous feeding in critically ill adult patients. The primary outcome was peak plasma insulin within three hours of an intermittent feed compared to continuous feeding.

## METHODS

### Ethics and Trial Registration

This trial was registered prospectively with a Clinical Trials Registry (clinicaltrials.gov - NCT06115044). We obtained ethical approval from the Wales Research Ethics Committee 3 prior to data collection (reference 23/WA/0297). A research-without-prior-consent model was used. This approach was deemed appropriate in patient and public consultation and approved by the Research Ethics Committee. Informed consent was sought once patients regained capacity. Further details are in the published trial protocol.(26)

### Study Design

This study was a prospective, parallel group, randomised, open-label trial. This was a single-centre study, conducted in a 48-bed mixed ICU, set within a 996-bed teaching hospital and major trauma centre.

### Participants

Adult patients anticipated to require gastric enteral nutrition for >48 hours were eligible to participate. Participants were excluded if any of the following applied: anticipated to be on enteral nutrition for <48 hours; requiring parenteral or jejunal nutrition; trophic feed only (e.g., lactate >4); deemed to be at high risk of refeeding syndrome; previous gastrointestinal surgery or pathology; diabetic emergency; pregnancy or prone positioning.

Recruitment took place between December 2023 and April 2024. Participants were screened for eligibility 7 days of the week and could be recruited within 24 hours of starting enteral nutrition by an appropriately trained healthcare professional on the study delegation log. Participants were monitored closely for the core study outcomes for 48 hours after study entry (trial intervention period), and length-of-stay outcomes were recorded until hospital discharge. The trial ended on completion of the follow-up of the final participant.

### Randomisation and Blinding

The randomisation sequence was generated using the National Cancer Institute Clinical Trial Randomisation Tool and placed into sealed opaque envelopes by an individual independent to the study prior to enrolment. Patients were randomised in a 1:1 ratio to the intermittent diurnal or continuous feeding trial arms by a member of the study team. Following randomisation, the study was open label, with only the study statistician blinded to group allocation. An open-label trial design was chosen due to both the difficulty of masking the pattern of feed administration, and to allow early identification of possible adverse effects.

### Trial Intervention

A detailed description of the trial intervention has been published previously.(26) In brief, the intervention was an adjustment in the delivery of gastric feed, to an intermittent diurnal pattern, for a period of 48 hours (Figure 1).

**Figure 1:**
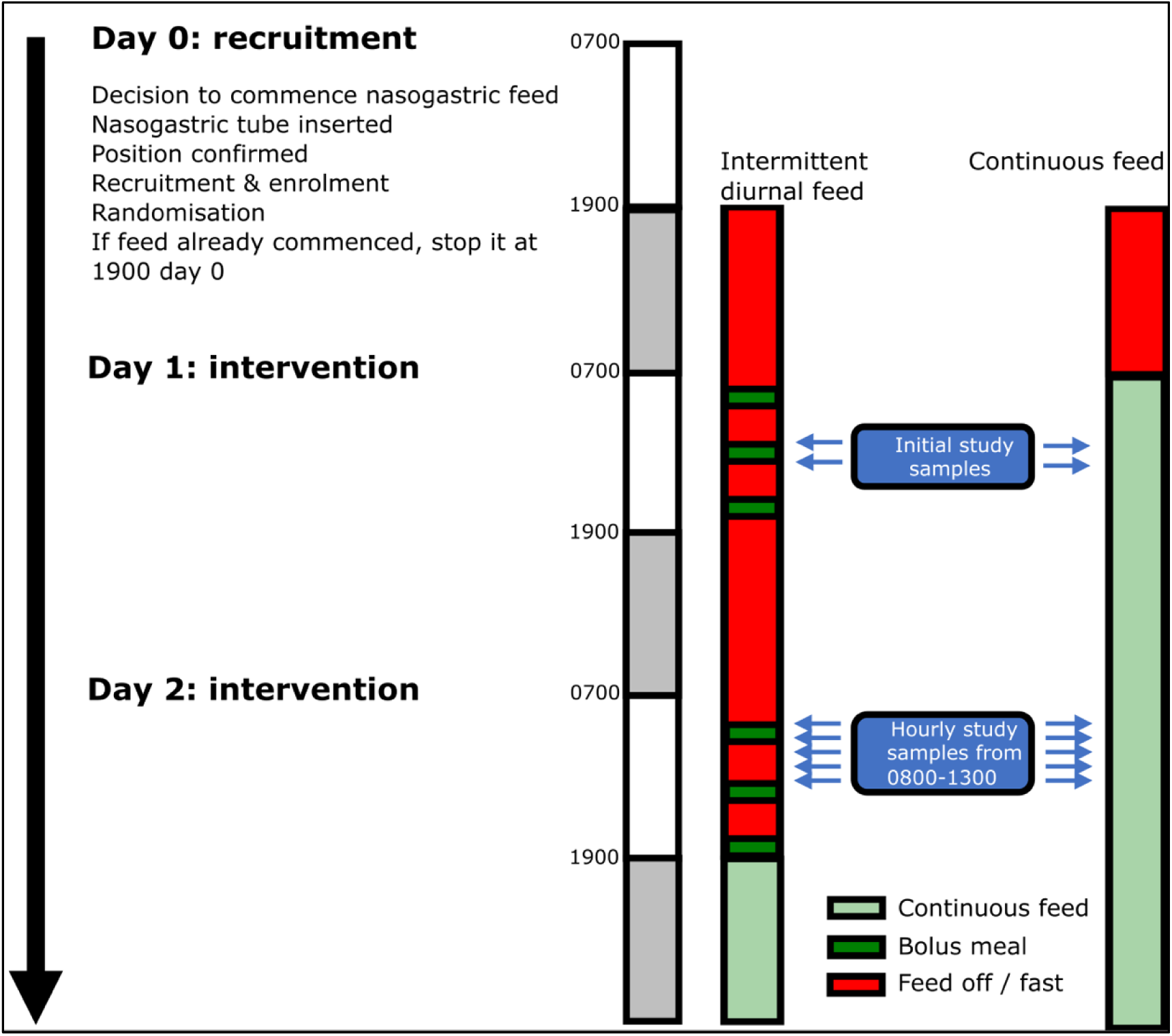
Feeding and sampling timelines.

The intermittent feeds (each one third of the calculated daily feed requirement) were administered at 8:00, 13:00 and 18:00 each day. The comparator group followed a continuous nasogastric feeding regime, as per local nutrition guidelines, to deliver the calculated daily feed requirement (see Supplementary Material). At the end of the 48-hour intervention period, patients reverted to the local continuous enteral feeding regimen.

### Outcomes

The primary outcome was peak plasma insulin reached within 3 hours of a feed compared to the peak over the equivalent time period in the continuous feed group. This was measured for the morning feed on day 2 of the study. Blood samples were taken hourly between 8:00 and 13:00 from an indwelling intra-venous/intra-arterial catheter to allow identification of any peak in plasma insulin levels (Figure 1).

Secondary outcomes included endocrine and metabolic measures and assessment of feasibility, tolerability and efficacy of the intervention. Planned endocrine and metabolic outcomes assayed from blood plasma included: c-peptide; glucose; ketones (beta-hydroxybutyrate); urea; non-esterified fatty acids; triglycerides and glycerol. Feasibility outcomes were: percentage of target nutrition achieved (per 24-hour period); absolute calories delivered and protocol compliance. Tolerability outcomes were episodes of vomiting/24-hour period; episodes of aspiration of feed; delayed gastric emptying (defined as gastric residual volume >250 mL×2 in a 24-hour period)(27); ileus; diarrhoea (passage of type 6 or 7 stool according to the Bristol Stool Chart or >3 stool/24 hours) and constipation (defined for this study as absence of bowel opening during the 48hr study period). (28) Indications of efficacy were based on ICU and hospital length of stay; ICU and hospital mortality; Delta-SOFA (Sequential Organ Failure Assessment) score between day 0 and day 2.

### Study Procedures

Participants were recruited on study day 0, initiating an overnight fast from 19:00 on that day, marking the start of the 48-hour intervention period. For the diurnal intermittent group, three feeds were administered each day via a volumetric pump over a period of 30-60 minutes. On study day 1, each feed consisted of 200ml (600ml/24 hours). On study day 2, each feed provided one third of the individual’s daily caloric requirements, determined using a weight-based equation set by intensive care dieticians in accordance with the local nutrition guideline (see Figure S1 in Supplementary Material).(8) Patients in the continuous feeding group had an identical feed amount calculated and delivered as a steady infusion over each 24 hours. Patients in the intermittent diurnal feeding group restarted the usual local continuous enteral feeding regimen at 12:00 on day 3 to prevent overfeeding. Patients were monitored within the study for an additional 12 hours after the 48-hour intervention period to identify any adverse effects potentially attributable to the intervention. The feed type used was Nutrison Protein Plus (Nutricia, UK).

The protocol paper provides further details on: concomitant medication; assessment of compliance; blood sampling and storage; analysis; participant study exit criteria; data collection/management/analysis and monitoring.(26)

### Biochemical analysis

Plasma insulin was quantified using an enzyme-linked immunosorbent assay (Mercodia, Sweden). Plasma glucose, non-esterified fatty acids (NEFA), glycerol, triglycerides, and urea were determined using commercially available spectrophotometric assays (Randox, UK). The assay for c-peptide failed quality control, therefore no results are available for this analyte.

### Statistical Analysis

The statistical analysis plan was published a priori in the protocol paper.(26) The desired sample size of 30 participants was based on findings from a study of healthy individuals, where the intermittently fed group had a mean (±SD) peak plasma insulin concentration of 373±204 pmol/L 2 hours after feeding, compared to 58±41 pmol/L in the continuously fed group.(29) This standardised effect size of 2.14 indicated that 6 participants per group would provide 90% power (p=0.05). Anticipating a smaller effect size in the critically ill population due to reduced feed volumes and impaired physiological capacity to mount a response, the target sample size was adjusted to detect an effect size of 1.26, requiring 11 participants per group for 80% power. To account for a 25% dropout rate, the sample size was increased to 30 participants.

Data were analysed by a blinded statistician using an intention-to-treat analysis set. Two-sided statistical tests were used throughout, p≤0.05 taken as statistically significant. The primary outcome was peak plasma insulin within 3 hours of the morning intermittent feed on the second study day, compared to continuously fed patients at the equivalent time point. A two-sample t-test was used to analyse the primary outcome, with effect sizes reported with 95% CIs. Similar tests (providing no violation of assumptions) were used to analyse the secondary hormonal and metabolic indicators.

Length-of-stay outcomes were compared using Kaplan-Meier analyses and the log-rank test. The percentage achieving target nutrition (per study day) was reported with 95% confidence interval for between-group differences.

## RESULTS

266 patients were screened between December 2023 and April 2024. Of these, 30 patients were enrolled; 13 patients were randomised to intermittent diurnal feeding and 17 to continuous feeding (Figure 2). In the intermittent diurnal feeding arm, one patient required jejunal feeding and therefore had to discontinue the study intervention, and one patient withdrew consent. The final analysis included 11 patients in the intermittent diurnal feeding group and 17 patients in the continuous feeding group.

**Figure 2:**
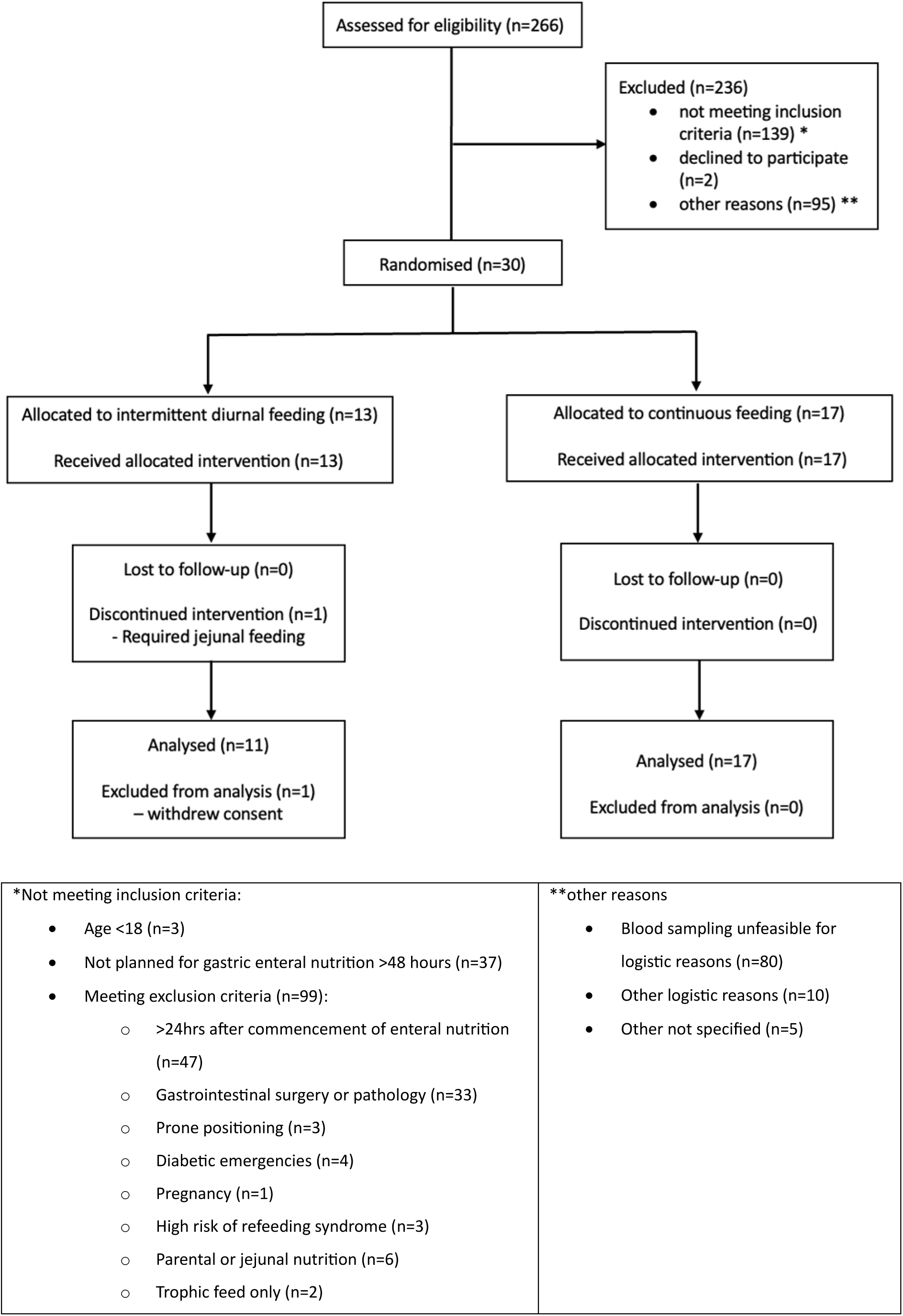
CONSORT diagram showing flow of participants through the trial.

Clinical and demographic characteristics were well matched at baseline (Table 1). There were 2 participants with diabetes in the continuous feeding group; both participants had diet-controlled type 2 diabetes and were not on any oral hypoglycaemic agents or insulin upon enrolment to the study.

**Table 1:** Baseline characteristics.

|  | **Intermittent diurnal feeding**<br>**(n=11)** | **Continuous feeding (n=17)** |
| --- | --- | --- |
| Sex (male) | 6/11 (54.5%) | 10/17 (58.8%) |
| Age (yrs) | 60 (54-70) | 60 (54-72) |
| Mass (kg) | 77 (65-83) | 75 (67-83) |
| Height (cm) | 174 (165-181.5) | 175 (165-182) |
| Ethnicity | 3/11 (27%) - English, Welsh,<br>Scottish, Northern Irish or<br>British<br>1/11 (9%) Black, Black<br>British, or Caribbean<br>background<br>7/11 (63%) – any other ethnic<br>group | 7/17 (41%) - English, Welsh,<br>Scottish, Northern Irish or<br>British<br>1/17 (6%) - Black, Black<br>British, or Caribbean<br>background<br>9/17 (53%) – any other ethnic<br>group |
| SOFA score | 7 (5.25-9) | 7 (6 – 9) |
| ICNARC score | 17.5 (14.25-21) | 17 (14.75 – 21.25) |
| APACHE II score | 13 (10.25 – 21.25) | 15 (10.5 – 22) |
| Presence/absence of<br>diabetes | 0 | 2/17 (both type 2) - 12% |
| Medication (insulin, oral<br>hypoglycaemic agents) | No insulin/hypoglycaemic<br>agents | No insulin/hypoglycaemic<br>agents |
| Time from admission to<br>ICU to start of enteral<br>feeding (hours) | 11.5 (9.8 – 31.4) | 15.3 (7.6 - 17.9) |
| Time from start of enteral<br>feeding to enrolment into<br>study (hours) | 23.3 (18.6 – 33.5) | 31.4 (22.9 - 35.3) |

Data are shown as median (IQR) or frequencies (n, (%)). APACHE II: Acute Physiologic Assessment and Chronic Health Evaluation; ICNARC: Intensive Care National Audit and Research Centre; ICU: intensive care unit; IQR: interquartile range; SOFA, Sequential Organ Failure Assessment.

Peak plasma insulin was significantly higher in the intermittent feeding group (295.1 ± 167.8pmol/L) versus continuous (128.1 ± 57.2pmol/L, p = 0.001) (Figure 3A), This is also shown by the greater maximum increase from baseline in the intermittent group (Figure 3B) with a clear peak noted 2 hours after the start of the intermittent feed (Figure 3C).

**Figure 3:**
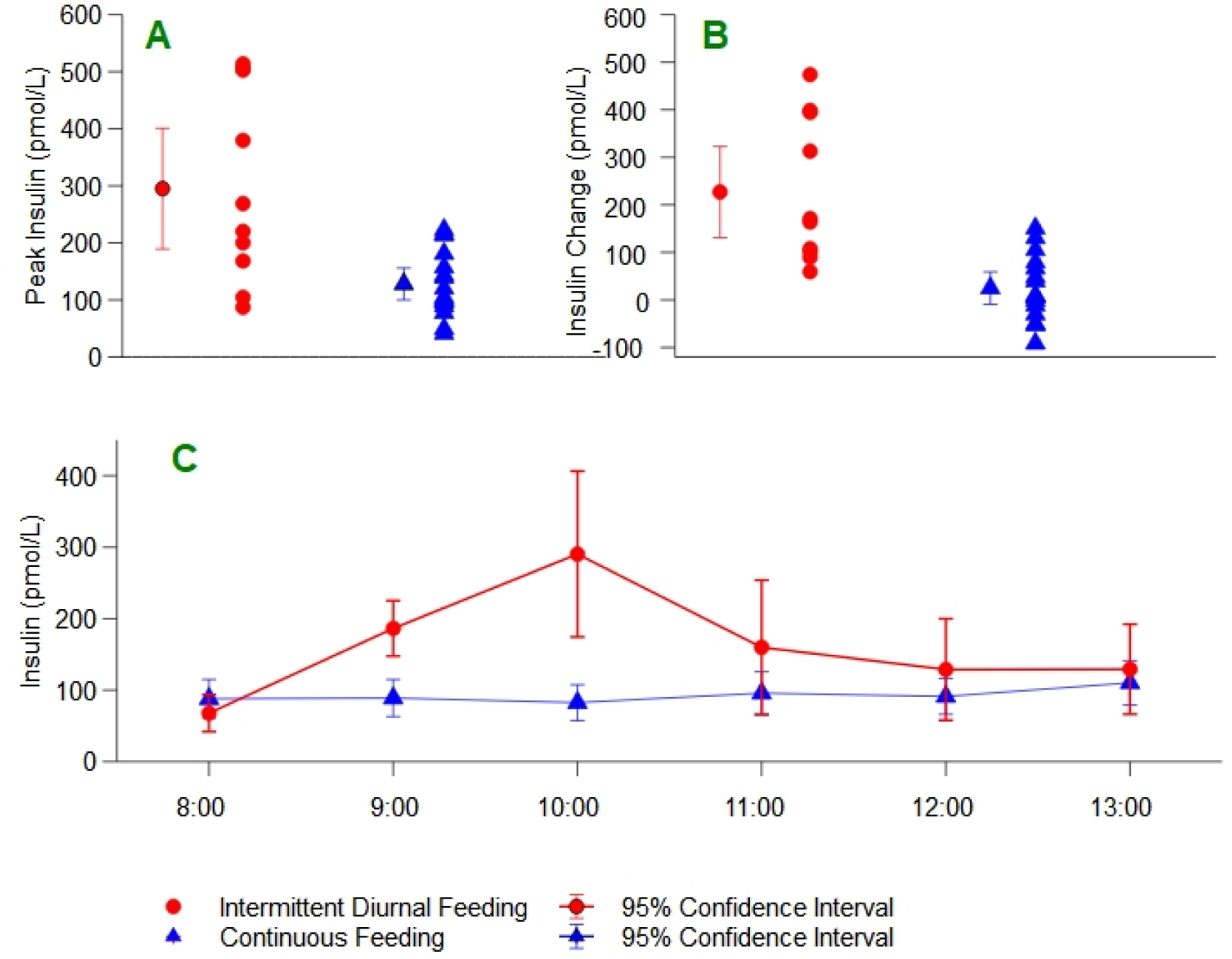
Plasma insulin. A: mean peak plasma insulin between 0800 and 1100 study day 2; B: mean increase in plasma insulin from 0800 baseline study day 2; C: Plasma insulin hourly during sampling period.

The difference between groups remained significant in a *post hoc* exploratory sensitivity analysis which excluded a single patient who was commenced on an exogenous insulin infusion (in intermittent diurnal feeding group, see Figure S2 in Supplementary Material). Plasma glucose concentrations during the sampling period were significantly elevated in the intermittent group at 10:00 (10.6 ± 2.5mmol/L, versus 8.4 ± 2.4mmol/L, p=0.04, Figure 4A). However, there were no episodes of hyper- or hypoglycaemia in either group and glucose variability, defined as the difference between the highest and lowest glucose recordings, was comparable between the two groups (Table 2).

**Figure 4:**
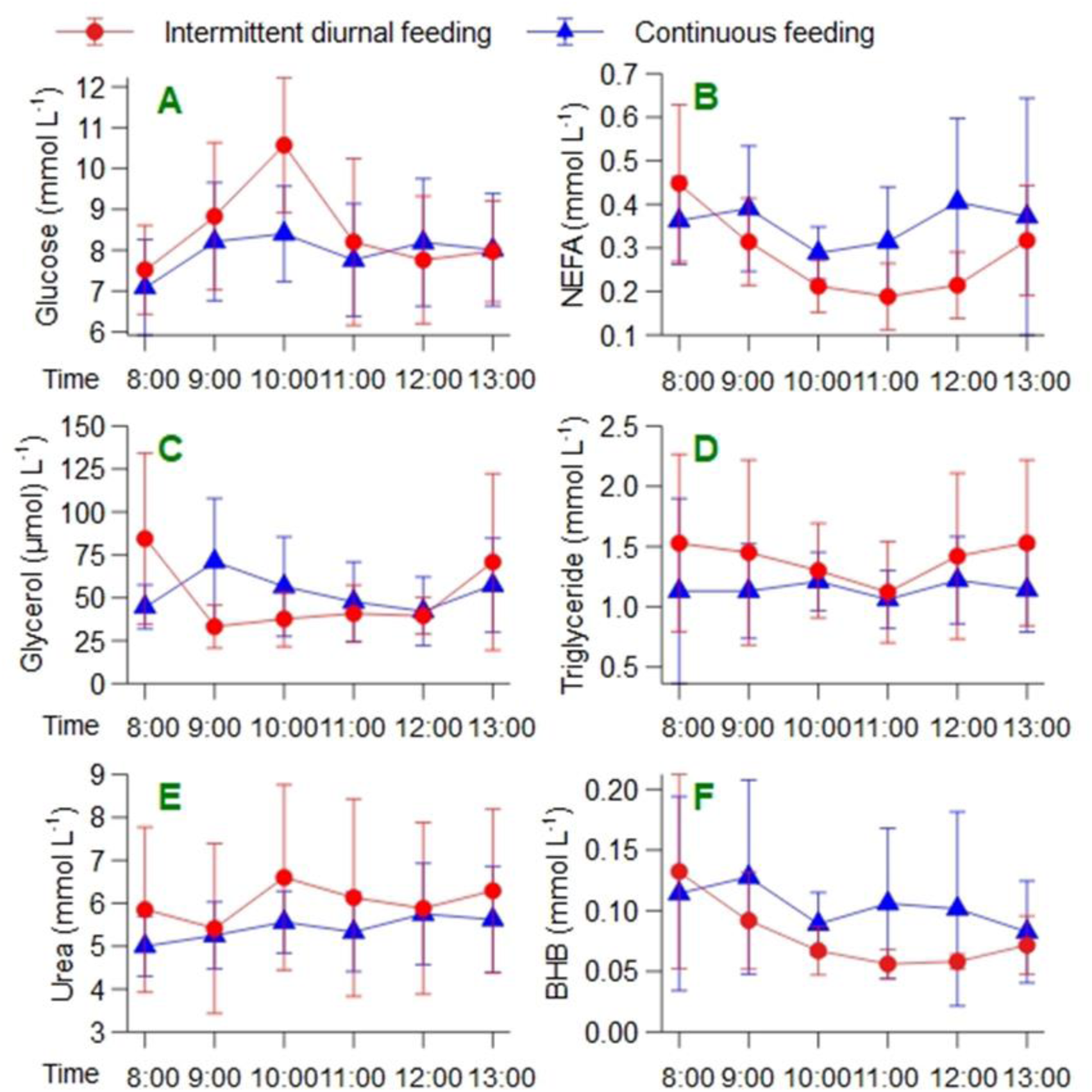
Comparison of metabolites. A) Glucose concentrations showed a significant rise at 10am in the intermittent group paralleling the peak in insulin (p=0.04). There was a non-significant pattern of a reduction in NEFA, glycerol and BHB concentrations across time in the intermittent group (B, C and F) without evidence of increases in triglyceride or urea concentrations (D and E).

**Table 2:**
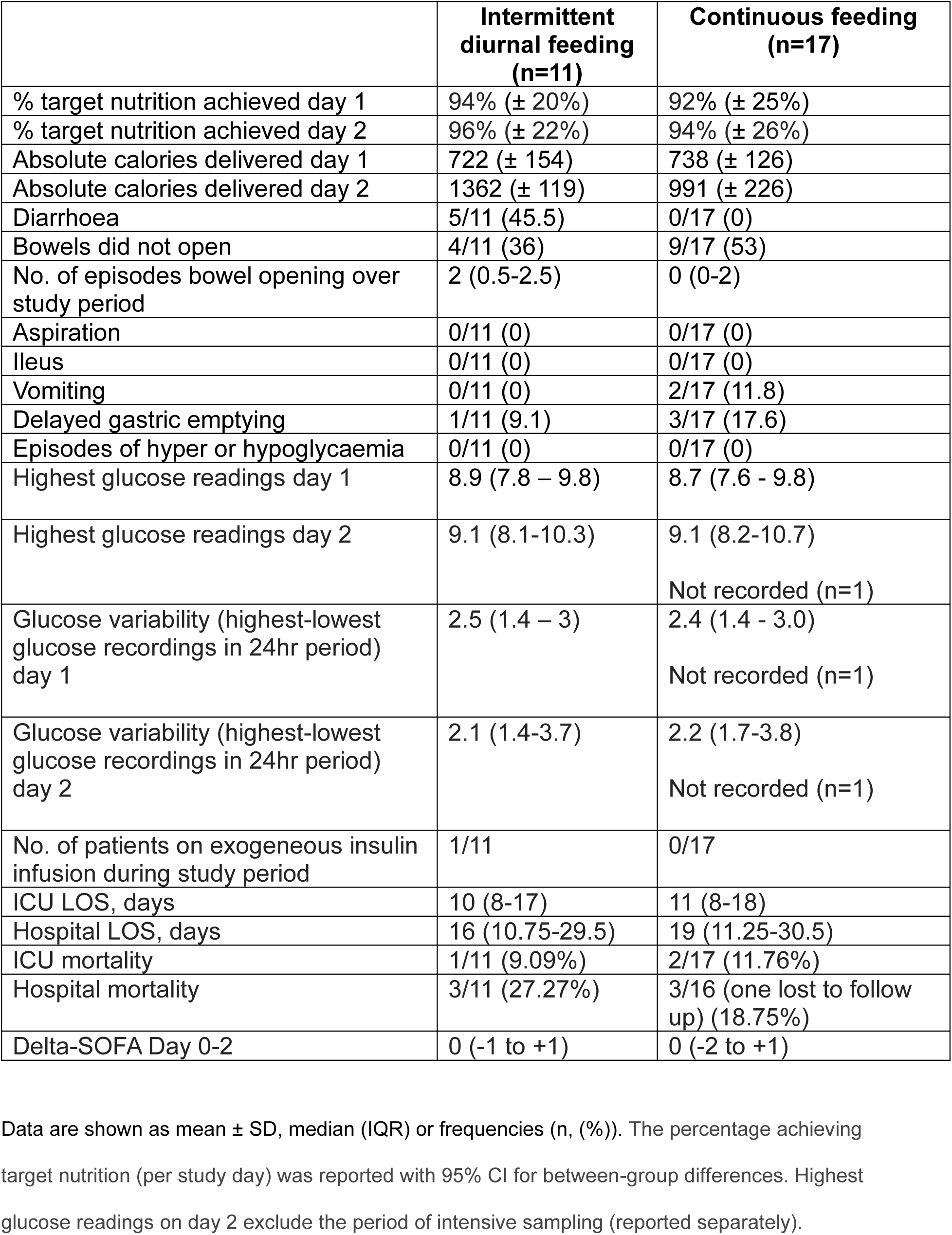
Secondary Outcomes.

A higher percentage of patients in the intermittent group opened their bowels (64%) compared to the continuous group (47%) during the 48-hour study period (Table 2). Patients in the intermittent feeding group had more frequent bowel movements over the 48-hours, with a median of 2 episodes (interquartile range 0.5-2.5), compared to the continuous feeding group, which had a median of zero episodes (interquartile range 0-2). Diarrhoea (loose or liquid stool) was more frequent in the intermittent feeding arm (5/11 patients) compared to continuous (0/17 patients) (p=0.005). Both groups met caloric targets during the study period and no adverse events were recorded. There was no increase in gastric stasis, vomiting, aspiration or ileus in the intermittent group (Table 2). There were no other statistically significant between-group differences in the analysis of the metabolites (Figure 4) and there was no significant difference between the groups in ICU or hospital length of stay (Supplementary figure 3).

## DISCUSSION

This study examined the feasibility and effects of a novel regimen of diurnal intermittent enteral feeding compared to continuous enteral feeding on hormonal and metabolic markers in a critically ill population. Our main finding is that intermittent diurnal nutrition preserves a postprandial insulin response in critically ill adults similar in timing and magnitude to that seen in healthy adult volunteers.(29) There is no corresponding increase in plasma glucose variability, nor in the incidence of hyper- or hypo-glycaemia. Other recent metabolic studies of intermittent feeding in critically ill adults did not report on the endogenous insulin response.(30,31)

The intermittent feeding regimen was shown to be feasible and provided comparable calorie delivery to standard continuous feeding without any increase in the incidence of delayed gastric emptying, vomiting, aspiration and with an indication that it may improve bowel motility. From an economic perspective there would be few costs associated with implementation, given the intervention only adjusts the timing of feed delivery. In this our findings are in line with those of other investigators, who also conclude that intermittent feeding is feasible and safe.(32)

### Insulin

Insulin is a key regulator of metabolism. An increase in insulin in response to feeding will promote cellular glucose uptake and use, along with skeletal muscle amino acid uptake and protein synthesis, with a net anabolic effect. There may also be other direct effects on vital organs such as brain, kidney and endothelium that are all important in critical illness.(33)

Additionally, the phasic nature of the insulin peaks and troughs are thought to be an intrinsic component of cell signalling that is crucial for usual homeostatic effects of insulin and that cannot be replicated with continuous feeding.(34) The insulin response to glucose is graded, with β-cells exhibiting a threshold-dependent secretion pattern.(35, 36) Acute increases in glucose levels, such as those seen with intermittent feeding, are more likely to trigger significant insulin release compared to the gradual nutrient delivery associated with continuous feeding, which may not reach the necessary threshold for substantial insulin secretion.

Following an overnight fast insulin concentrations are low which permits increased rates of lipolysis in adipose tissue to provide circulating fatty acids as fuel.(37) This is typically reflected by high circulating concentrations of glycerol and non-esterified fatty acids. In the postprandial state, increased insulin concentrations contribute to controlling glucose concentrations both directly (via suppression of endogenous glucose production and stimulation of peripheral glucose uptake) and indirectly, via suppression adipose tissue lipolysis and thereby reduced competition for mitochondrial substrate oxidation between glucose and fatty acids.(38) The insulin induced suppression of lipolysis also reduces substrate availability for ketogenesis.(39) The overall picture of the hormonal and metabolic patterns observed in the intermittent group in this study (low insulin, with high glycerol and fatty acid concentrations in the fasted state, and suppressed glycerol, non-esterified fatty acid and beta-hydroxybutyrate concentrations in the fed state, without evidence of increases in triglycerides or urea concentrations) appear consistent with this, and may lend support to the argument that this intermittent feeding regimen preserves a more “normal” metabolic and hormonal pattern than does continuous feeding in critically ill patients. Whether this holds for longer periods of intermittent feeding and in larger groups of patients, and whether any difference translates into clinical benefit, are questions that remain to be answered.

### Tolerability

This intermittent diurnal feeding intervention was well tolerated. Although we noted a peak in plasma glucose after an intermittent feed, there was no additional risk of hyper- or hypoglycaemia and no increase in the glucose variability, likely reflecting the increased secretion of insulin and other gut hormones promoting glucose uptake and utilisation. One previous meta-analysis highlighted an increased risk of gastrointestinal intolerance and aspiration compared to continuous feeding (40), but this was not observed in our study where there was no increase in gastric stasis, vomiting, aspiration or ileus. We observed an increased incidence of diarrhoea in the intermittent diurnal feeding arm. We defined diarrhoea as passage of type 6 or 7 stool according to the Bristol Stool Chart, or >3 stool/24 hours – all of our patient’s diarrhoea diagnoses were on the basis of the presence of loose stool rather than frequency. Indeed, some studies define diarrhoea simply as the passage of >5 stools per day,(41) so by this criterion our result represents an overestimate of the incidence of diarrhoea. Given that constipation is a problem for over half of critically ill patients in the first week of ICU admission, intermittent feeding may actually represent maintenance or restoration of normal gut motility, given the higher rates of bowels opening in the intermittent feeding group.(42)

### Generalisability and sources of bias

Our study has several limitations. Inherent to the design is a lack of blinding that may introduce bias, although this is mitigated somewhat by the objective nature of the primary and secondary outcomes. Blinding was maintained for both the plasma metabolite/hormonal assays and the statistical analysis of all measures. In a definitive study it may be possible to maintain blinding of the study teams by having pre-programmed pump settings and concealment of the displayed rates, although that would compromise patient mobility benefits. In addition, this was a single centre study. Feeding guidelines and practices may vary between Intensive Care Units, as may case mix. However, given the primary outcome is measuring a fundamental physiological response we suggest that the findings are likely to be generalisable and justify further investigation in multiple centres. The sample size is small and the period of intervention short reflecting the early-phase nature of the study but was calculated prospectively against a key hormonal outcome (pulsatile insulin release) with allowance for both a smaller effect size and for drop out.

### Specific trial limitations

We were unable to account for non-nutritional calories, including the contribution of lipid-soluble medications or intravenous dextrose. Although theoretically balanced between groups due to randomisation, this may be worth measuring as a potential confounding variable in future large-scale studies. To account for the loss of c-peptide, we ran a post-hoc sensitivity analysis excluding the only patient on exogeneous insulin (in the intermittent diurnal group), which made no difference to the primary outcome.

By chance the study only included one patient with diabetes, who was managed with diet alone. Whether the insulin response to intermittent feeding would be similar in critically ill diabetic patients, and whether there would be any difference between type 1 and type 2 diabetes, will require further investigation.

To avoid harm to the participants from unnecessary oversampling, and due to the resource-intensive nature of blood sampling; we chose to focus intensive blood sampling around the morning feed on study day 2. As a result, there is a chance that we have failed to detect other metabolic phenomena that may have occurred during the study period. We also are unable to comment on any metabolic changes that occur after the study period. Nevertheless, demonstrating an insulin response to intermittent feeding that is both different to that seen with continuous feeding and similar to that in health is consistent with a genuine metabolic phenomenon that has the potential to influence the course of critical illness.

## CONCLUSION

Diurnal intermittent feeding, compared to continuous feeding, preserves the endogenous insulin response and is consistent with a more physiological metabolic milieu in critically ill adults, without additional risk of dysglycaemia. This was an efficacious and well-tolerated intervention in the ICU which offers other pragmatic benefits, such as improved bowel motility and fewer restrictions on patient mobility. Further research is needed to assess other clinically important outcomes including on long-term survival and function. Future work should also address acceptability of intermittent feeding to patients and professionals and consideration of the cost-effectiveness of the intervention.

## Data Availability

All data produced in the present study are available upon reasonable request to the authors

## Abbreviations

ICU: Intensive Care Unit
LOS: length of stay
NEFA: non-esterified fatty acids
NG: nasogastric
PPI: Public and Patient Involvement
SOFA: Sequential Organ Failure Assessment
SD: Standard Deviation

## Author Contribution

See Author Statement File.

